# Immunochromatographic assays for COVID-19 epidemiological screening: our experience

**DOI:** 10.1101/2020.05.28.20116046

**Authors:** Andrea Bartolini, Margherita Scapaticci, Marina Bioli, Tiziana Lazzarotto, Maria Carla Re, Rita Mancini

## Abstract

In March 2020, the World Health Organization (WHO) declared a pandemic caused by severe acute respiratory syndrome coronavirus 2 (SARS-CoV-2). Due to the absence of effective treatment or biomedical prevention, understanding potential post infection immunity has important implications for epidemiologic assessments. For this reason, increasing number of in vitro diagnostic companies are developing serological assays to detect antibodies against SARS-CoV-2, but most of them lack the validation by third parties in relation to their quality, limiting their usefulness. We submitted to serological screening by two different immunochromatographic (IC) rapid testing for detection of IgG and IgM against SARS-CoV-2, 151 asymptomatic or minimally symptomatic healthcare workers previously tested positive for SARS-CoV-2 RT-PCR in order to evaluate the performance of rapid assays. Results showed discrepancies between molecular and IC results, and an inconsistency of immunoglobulins positivity patterns when compared to ELISA/CLIA results, highlighting the absolute necessity of assays performance validation before their marketing and use, in order to avoid errors in the results evaluation at both clinical and epidemiological level.

## Introduction

Coronavirus disease 2019 (COVID-19) is an infectious disease caused by severe acute respiratory syndrome coronavirus-2 (SARS-CoV-2), which first appeared in Wuhan, China, in December 2019 and then quickly spread worldwide, so that the outbreak was declared a pandemic in March 2020 by the World Health Organization (WHO). The responsible pathogen of COVID-19 is a positive-sense single-stranded RNA virus, belonging to the Coronaviridae family (1). Because no vaccine and not approved specific treatment are currently available (2, 3), even if many trials are in progress, WHO has defined mandatory to mobilize research to control the COVID-19 emergency. So, epidemiological studies, clinical characterization and management, infection prevention and control, including health care workers’ protection, have a crucial role to quickly identify sick people, treat them, and better estimate how widely the virus has spread (4). Knowledge of diagnostic tests for SARS-CoV-2 is still evolving and the immune response against the virus is not yet fully understood. Nevertheless, as for other well-known infectious diseases, the host immunity certainly has a role in fighting against the pathogen and in predicting the prognosis (5). Real-time reverse-transcript Polymerase Chain Reaction (real-time RT-PCR), performed using nasopharyngeal swabs or other upper respiratory tract specimens (i.e. throat swabs, or salivary samples) is the commonly used test for diagnosis of COVID-19 (6), but even if widely adopted as the gold standard method, it presents some limitations (7). False-negative results can occur due to poor quality of the specimen collected, for example when the sample is collected with an inappropriate timing in relation to the illness onset, or to an inappropriate specimen handling or shipping to the Laboratory Department. At the same time, false positive results can be due to contamination or technical errors. Unlike molecular testing, detection of an immune response to the virus is an indirect marker of infection and the development of robust serological tests and their appropriate utilization are essential to support ongoing public health effort to effective epidemic prevention (8). Serological tests could be used for epidemiologic studies on a large scale population, especially in asymptomatic population or those presenting mild symptoms (including healthcare workers) (5) to theoretically determine the number of people exposed to the virus in a population and to reveal who have had the disease and are currently immune. However, up to date, there are only few data published about their efficiency, often controversial (9–11).

An immunoglobulin test measures the level of antibodies produced and secreted by B lymphocytes when the immune system tries to protect the body from bacteria, viruses, etc. Immunoglobulin M (IgM) is the first antibody the body makes when it fights a new infection. Then, Immunoglobulin G (IgG) starts to be produced to ensure a robust, longer lasting immunity (7). Literature data report that IgM and IgG against SARS-CoV start to be detected after 3–7 days from the onset of the infection and persist for a period of 2–3 years (12–15), similarly the first studies about the antibodies against SARS-CoV-2 show that they started to be detected after 4–5 days (7, 16).

Different assays can be used for SARS-CoV-2 antibodies revelation:

– immunochromatographic rapid test (IC)
– enzyme-linked immunosorbent assay (ELISA)
– chemiluminescent immunoassay (CLIA).

From 3^rd^ to 27^th^ of April 2020, 10,945 asymptomatic or minimally symptomatic healthcare workers were submitted to qualitative serological tests for SARS-CoV-2 IgM and IgG during the COVID-19 medical surveillance at the Local Health Unit of Bologna in order to determine their immunological state. Due the contacts caused by the type of work done, some of them were considered much more exposed to risk of infection, for this reason they had previously been submitted to RT-PCR, so that 151 individuals already tested positive for SARS-CoV-2 RT-PCR (nasopharyngeal or oropharyngeal samples). Our aim was to evaluate the performances of two different IC assays, submitting to serological testing the 151 healthcare workers previously tested positive for SARSCoV-2 RT-PCR and trying to find a correlation between the molecular method, that is considered the gold standard and rapid IC tests actually available. Finally, we submitted the samples who tested positive by IC to ELISA/CLIA assays for a further evaluation.

## Methods

Health professional workers samples were collected into collection tube containing EDTA by venepuncture and the plasma was immediately separated by centrifugation to be submitted to qualitative rapid serological tests.

Due to the emergency, our availability of IC testing was affected from difficulties to find the same kit continuously, so we were forced to use two different rapid tests for two different groups of individuals. During the first part of the surveillance we used the KHB® Diagnostic Kit for SARSCoV-2 IgM/IgG Antibody (Colloidal Gold), while for the second part we used Cellex qSARS-CoV-2 IgG/IgM Cassette Rapid Test. Both the methods were characterized by a lateral flow chromatographic immunoassay in a cassette format. When an adequate volume of test specimen, according to manufacturer instructions, is dispensed into the sample well, the specimen migrates by capillary action along the cassette. Immediately after the specimen, two or three drops of the sample diluent are added to the same area. The cassette is incubated for 15 minutes at room temperature then it is possible to read the result. IgG and/or IgM anti SARS-CoV-2, if present, will bind to the SARS-CoV-2 conjugate constituting an immunocomplex that can be captured by the anti-human IgG and/or IgM conjugate. The presence of the captured immunocomplex is visible due to its precipitation in a coloured red band. If the specific antibodies IgG/IgM are absent or are present in extremely low concentrations to be detected, no test line will appear except for the internal control line. The samples that showed presence of IgG and/or IgM by rapid testing were transferred to Microbiology Department and then subjected to iFlash-SARS-CoV-2 IgG/IgM assay (CLIA) supplied by Pantec S.r.l. and/or Anti-SARS-CoV-2 IgG ELISA assay supplied by EUROIMMUN ITALIA S.r.l. for a further determination of IgG and IgM antibody against SARS-CoV-2. The remaining plasma were stored at –20°C for further analysis.

### Ethical approval statement

Sampling was conducted within the surveillance program established by the Emilia-Romagna Region and did not require ethical approval (17). The samples were collected after informed consent was given.

### Statistical analysis

Statistical significance was assessed using Fisher’s exact test and a *p*-value of <0.05 was considered significant.

## Results

As showed in table n. 1 we stratified the results according to the time intercourse between the positive RT-PCR and the IC testing, without distinguishing between IgG and/or IgM positivity.

**Table 1.** Results of IC testing according to the time intercourse between the SARS-CoV-2 RT-PCR positive. KHB/Cellex positive include the following patterns: IgG+/IgM−, IgG−/IgM−, IgG+/IgM+

| Days after RT-PCR positivity | KHB positive | KHB negative | Cellex positive | Cellex negative |
| --- | --- | --- | --- | --- |
| 0-5 | 1 | 1 | 1 | - |
| 6-10 | 3 | - | - | - |
| 11-15 | 3 | - | 9 | 4 |
| 16-20 | 2 | 4 | 18 | 4 |
| 21-25 | 4 | 15 | 17 | 7 |
| 26-30 | - | 2 | 13 | 4 |
| 31-35 | - | - | 13 | 12 |
| 36-40 | - | - | 5 | 6 |
| 41-45 | - | - | 1 | 2 |

Only 90 out of 151 (59.6%) samples with the molecular positive result came out with at least one (IgM and/or IgG) positivity by IC assays moreover, comparing the two IC methods, we found a significant difference between the them (*p*=0.003). Moreover, calculating the proportion of individuals showing the presence of Anti-SARS-CoV-2 (IgM and/or IgG) by IC after the RT-PCR positivity, KHB showed a sensitivity of 35.3%, so much lower than Cellex, that even showed a poor sensitivity (66.4%).

In order to eliminate the possible bias due to different tests used in different periods for the calculation of the *p*-value, we tried to compare the two IC assays at the same period after the positive molecular result reducing the period of comparison: first we considered the tests carried out from day 0 to day 25 after positive RT-PCR and then, because literature data on SARS-CoV show that the development of antibodies generally reach the peak after 7–10 days from the onset of the infection (12–15), we further reduced the evaluation period considering the IC results obtained between 10–25 days after RT-PCR positivity. In both cases we had a significant difference (*p* < 0.001) between used IC methods, with a sensitivity of 39.4% for KHB and 75% for Cellex when the period of evaluation was 0 to 25 days, and a sensitivity of 32.1% and 74.6% for Cellex when the considered period was reduced from day 10 to day 25 after RT-PCR positivity. Even if both methods did not show a great efficiency in term of sensitivity, especially if we consider that the assays used for screening and epidemiological purposes need to have a high sensitivity, Cellex IC test seems certainly to have the best performances.

In table n.2 are showed the specific IgG and/or IgM positivity patterns obtained by IC tests. We observed that by Cellex IC assay we found 75 samples with both IgM and IgG positivity, most of them (90.7%) relieved between 11 and 35 days after RT-PCR positive results, with a peak between day16 and 25(46.7%), and only one sample IgG+ and one IgM+. On the other hand, KHB system did not seem to detect IgM at all regardless of the test period, in fact we had no sample with IgM only, eleven samples with IgG and only two samples IgM+/IgG+.

**Table 2.** Pattern of samples tested positive by IC

| Days after RT-PCR positivity | KHB |  |  | Cellex |  |  |
| --- | --- | --- | --- | --- | --- | --- |
|  | IgG+/IgM- | IgG-/IgM+ | IgG+/IgM+ | IgG+/IgM- | IgG-/IgM+ | IgG+/IgM+ |
| 0-5 | 1 | - | - | - | - | 1 |
| 6-10 | 3 | - | - | - | - | - |
| 11-15 | 3 | - | - | - | - | 9 |
| 16-20 | 1 | - | 1 | - | - | 18 |
| 21-25 | 3 | - | 1 | - | - | 17 |
| 26-30 | - | - | - | - | - | 13 |
| 31-35 | - | - | - | 1 | 1 | 11 |
| 36-40 | - | - | - | - | - | 5 |
| 41-45 | - | - | - | - | - | 1 |

Finally, 79 out of 90 samples showing a positivity by both RT-PCR and IC testing, were sent to Microbiological Department for ELISA/CLIA testing and 78 of them (98.7%) gave positive results (concordance of IgG/IgM patterns relieved by IC vs ELISA/CLIA is reported in table 3a for KHB system and table 3b for Cellex). Overall percentages of concordant patterns between each IC testing results and ELISA/CLIA were 15% for KHB and 43% for Cellex, proving a better performance of the latter. Anyway we noticed that KHB system showed a concordance with ELISA/CLIA only for two IgM+/IgG+ samples (15.4% of the total) found between 16 and 25 days after RT-PCR, while Cellex IC assay showed 28 completely concordant pattern compared to ELISA/CLIA (27 IgM+/IgG+ and one IgG+/IgM-), but 37 out of 75 IgG+/IgM+ patterns detected by Cellex showed a IgG+/IgM- pattern by ELISA/CLIA, suggesting that there is an overestimation of IgM by Cellex system or a their underestimation by ELISA/CLIA.

**Table 3.**
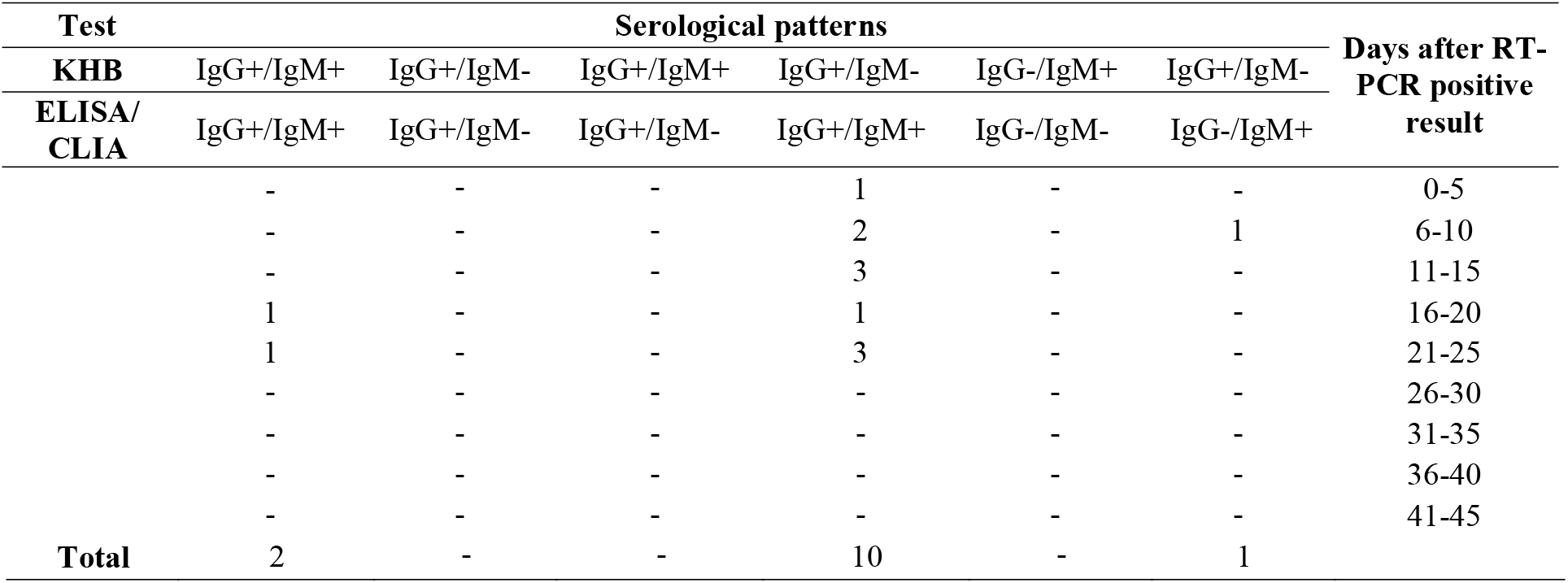
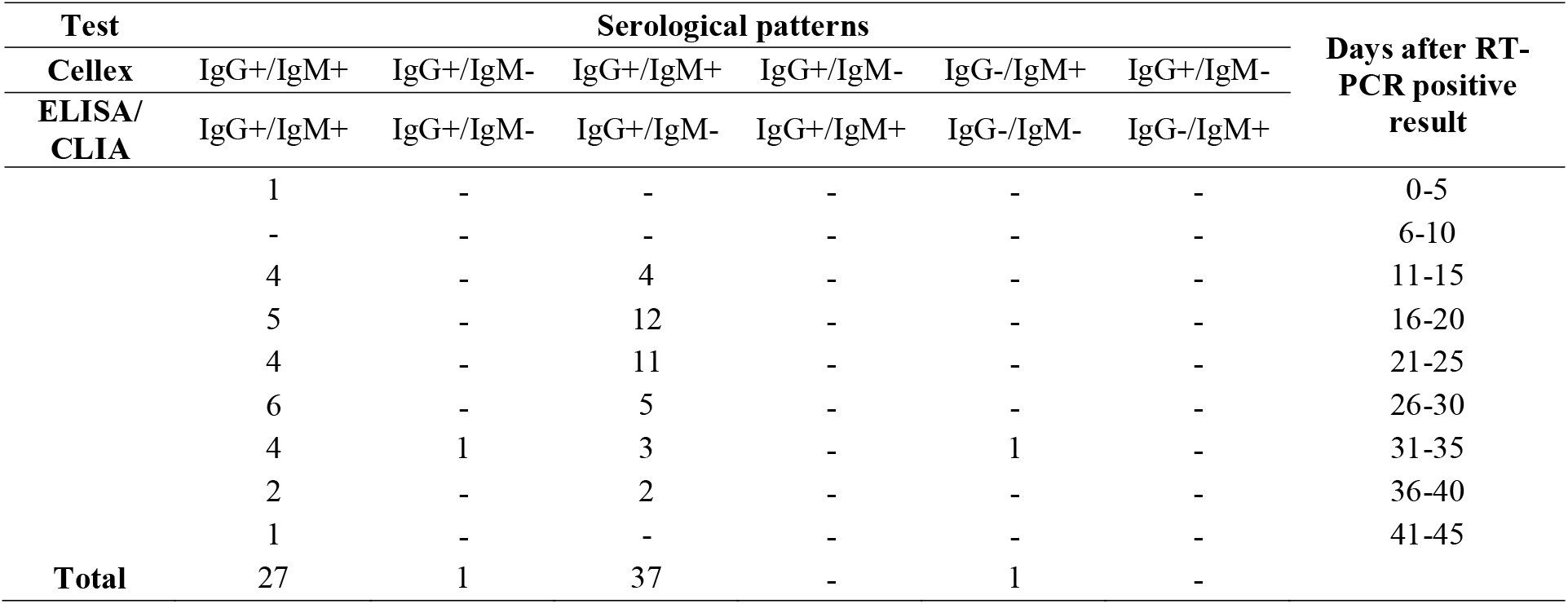
Concordance of IgG/IgM patterns relieved by KHC vs ELISA/CLIA

## Discussion and Conclusion

Given the acute and rapid onset of COVID-19, molecular testing of respiratory tract samples to detect SARS-CoV-2 RNA remains the preferred diagnostic test for assessment of symptomatic patients (2). Nevertheless, in addition to molecular testing, serologic assays able to detect antibodies against SARS-CoV-2 may help to reveal the immune response to the virus representing an indirect marker of infection. Development of robust serologic tests and their appropriate utilization and interpretation could be essential to maintain safe patient care standards and support ongoing public health efforts (8). Since it is not known how long immunization lasts and how effective it is against a new infection, appropriate epidemiological studies are needed. Detectable IgM and IgG antibodies against SARS-CoV-2 seem to develop within days to weeks of symptom onset in most infected individuals (9, 18), but why some patients seem not to develop a humoral immune response is uncertain, as well as is still unclear the relationship between antibody response and clinical improvement (19). For these reasons, reliable test data are necessary to better assess the validity of the information related to the pandemic spread, in term of positive cases, deaths, and immune individuals (12–14, 20). At the same time, the apparent lack of systematic and standardized protocols in most countries represents the main source of discrepancies in mortality rates. In our study we underline the necessity of using on a large scale validated testing in order to guarantee their usefulness in the epidemiological field, especially in emergencies such as this pandemic COVID-19, where the lack of sufficient time prevents the correct validation of the methods before their use. Moreover, we must consider that cross-reactivity may occur due to the structural similarity and the genomic sequence conservation of the immunogenic proteins of CoV species, that share common structural epitopes. For this reason, it is also important to know the epitopes to which the tests are directed (i.e. N protein-based serological assays, S protein-based assays, etc) (5). Our findings showed a great discrepancy between molecular and IC results, probably due to the poor sensitivity of the IC assays. Also, we found an inconsistency of immunoglobulins positivity patterns when we compared IC to ELISA/CLIA results, highlighting the absolute necessity of assays performance validation before their marketing and use, to avoid errors in the results evaluation at both clinical and epidemiological level. As recently confirmed by the circular of Italian Ministry of Health the qualitative rapid tests to SARS-CoV-2 antibodies detection need the validation by third parties in relation to their quality before being chosen for their clinical utilization and, to the best of the knowledge available today, there is currently no evidence produced by third parties about that (21). After all, because the methods are still in development, it is too early to make an evaluation of their usefulness in current epidemiology purposes (22). A limitation of our study was that we were not able to calculate the specificity of both IC assays, due to the lack of data about healthcare professional workers with RT-PCR negative submitted to serological evaluation.

## Data Availability

All data referred in the manuscript are available.

